# Tracing Tuberculosis Patients Lost to Follow-up: A Mixed-Methods Study of Community Health Promoter–Led Tracing in Kenya

**DOI:** 10.64898/2026.07.28.26359180

**Authors:** Moses M. Ngari, Stevenson K. Chea, Christine M. Mwamsidu, Deche Sanga, Geoffrey Katana, Fredrick M. Thuva, George U. Onyango, Catherine Kamau, Osman Abdullahi

**Affiliations:** KEMRI/Wellcome Trust Research Programme, Kilifi, Kenya; Department of Public Health, School of Health & Human Sciences, Pwani University, Kilifi, Kenya; Department of Nursing Sciences, School of Health & Human Sciences, Pwani University, Kilifi, Kenya; Global Fund AMREF Health Africa - Kenya; Kilifi County TB Control Program; Brain & Mind Institute, Agha Khan University - Nairobi, Kenya; JIVIS, Kilifi, Kenya

**Keywords:** Tuberculosis, Lost-to-follow-up, Community Health Promoters, Patient tracing, Treatment adherence, Kenya

## Abstract

Tuberculosis (TB) remains a major global public health threat, with an estimated 10.7 million incident cases and 1.23 million deaths in 2024. Treatment interruption undermines TB control efforts, yet the effectiveness of Community Health Promoters (CHPs) in tracing patients lost to follow-up (LTFU) is poorly understood. We assessed the proportion of LTFU TB patients successfully traced through CHP-led tracing strategy and explored challenges of implementation CHP strategy. We conducted a mixed-methods cross-sectional study in Kilifi County, Kenya, among adult TB patients recorded as LTFU between July 2022 and December 2023. Patients identified through the national electronic TB surveillance system were linked to their respective community health units and assigned to CHPs for tracing. CHPs attempted to contact patients by telephone and through repeated household visits and, where necessary, sought information from relatives, neighbours, and local administrative leaders. For successfully traced patients, quantitative data were collected using structured questionnaires. Qualitative data were collected through five focus group discussions with 30 traced TB patients and 30 in-depth interviews with CHPs, TB clinic staff, programme managers, and County Health Management Team members. Qualitative data were coded and analysed thematically using a primarily deductive approach guided by the study objectives, while allowing additional themes to emerge. Of 140 patients identified for tracing, 36 (25.7%, 95% confidence interval 18.7–33.8) were successfully traced; 90 (64.3%) had relocated outside Kilifi County and 14 (10.0%) had died. Qualitative findings indicated that treatment interruption was driven by financial hardship, including transport costs, medication side effects, stigma, and difficulties maintaining continuity of care after relocation. CHPs supported treatment adherence through health education, counselling, appointment reminders, and household follow-up. However, their ability to trace patients was constrained by patient mobility, limited financial support for community-based activities, shortages of health workers, competing responsibilities, and gaps in systems for tracking patients across administrative boundaries. CHP-led tracing reached only one-quarter of LTFU TB patients, largely because many had relocated outside the county. Health systems should strengthen patient tracking across county boundaries and equip CHPs with the resources, staffing, and support needed for effective community-based TB tracing.

## Background

Despite being preventable and curable, tuberculosis (TB) continues to rank among the leading causes of death worldwide, causing 10.7 million new cases and 1.23 million deaths in 2024 amid slowing progress towards global targets [1–3]. TB remains predominantly a disease of low- and middle-income countries, which account for approximately 98–99% of the global burden in terms of cases and deaths, with Africa disproportionately affected[4]. Kenya is among the high TB burden countries globally, with ongoing challenges in TB prevention, diagnosis, treatment, and retention in care[2]. Urgent actions are needed to end the TB epidemic, a goal adopted by World Health Organization (WHO) and United Nation member states[5, 6]. The End TB Strategy set ambitious targets to reduce TB incidence rate and mortality between 2015 and 2025. However, progress has been slow: by 2023, global TB incidence had declined by only 8.3% since 2015, far below the target of a 50% reduction by 2025, while TB-related deaths declined by 23% compared with the targeted 75% reduction over the same period[2, 3].

Loss to follow-up (LTFU) during TB treatment remains a major barrier to achieving optimal treatment outcomes and TB control. It leads to disruption in TB treatment which increases the risk of treatment failure, ongoing transmission, mortality, and the emergence of drug-resistant TB[7, 8]. Drug resistant TB is more expensive to treat, requires longer and more complex regimens and is associated with poor treatment outcomes and mortality[9]. The burden of LTFU varies widely across settings and within countries. In sub-Saharan region, only 79% of patients with drug-susceptible TB successfully complete treatment, with LTFU accounting for nearly half (47%) of all unsuccessful treatment outcomes[10]. The proportion of LTFU ranges from 6.3% in rural Kenya[11] and 13% in Urban informal settlement in Kenya[12] and in China[13], to 16.6% in Ethiopia[14], 18.1% in Brazil[15] and as high as 44.9% in Mozambique[16]. These variations reflect differences in health system capacity, socioeconomic conditions, and the availability of community-based support mechanisms.

Community Health workers (CHWs) now known as Community Health Promoters (CHPs) in Kenya, play a critical role as the first point of contact between communities and the health system[17, 18]. CHPs are embedded within the communities they serve and provide health education, basic preventive and promotive services, and linkage to formal healthcare. Although their roles are not explicitly defined, their proximity to households makes them particularly important for reaching marginalized populations, including those in rural areas, urban informal settlements, and settings with shortages of trained health professionals [19, 20]. CHPs have demonstrated effectiveness in supporting conditions that require sustained engagement with health system, such as maternal and child health, HIV, malnutrition, non-communicable diseases, and sickle cell disease [18, 21, 22]. However, CHPs face many challenges including inadequate training, limited supervision, lack of remuneration, and unclear scope of work, leading to high workload and reduced effectiveness [23, 24].

While TB treatment requires a minimum of six months of uninterrupted therapy, the specific role of CHPs in supporting and tracing TB patients lost to follow-up has not been well studied. Most previous studies have largely focused on CHP involvement in maternal and child health and HIV programs, areas that have historically attracted greater funding and programmatic attention [18, 22, 25–27]. Evidence on how CHPs can support TB programs, especially in tracing patients who default from care, is limited. Understanding the feasibility and challenges of CHP-led tracing is critical for informing community-based strategies to improve TB treatment retention. This study therefore aimed to assess the proportion of TB patients lost to follow-up who could be successfully traced by CHPs and to explore the challenges experienced by CHPs in supporting TB patients.

## Materials and Methods

### Study design

This was a mixed-methods cross-sectional study with both quantitative and qualitative components.

### Study settings

The study was conducted in Kilifi County, one of Kenya’s 47 counties, located along the country’s coastal region from 20^th^ April to 10^th^ June 2024. According to the 2019 national census, the county has a population of approximately 1.5 million people and a population density of 116 persons per square kilometre. The local economy is largely driven by tourism and fishing, reflecting its proximity to the Indian Ocean. Health services are delivered through 235 health facilities, including one county referral hospital: Kilifi County Referral Hospital.

TB diagnosis and management in Kilifi follow the Kenya National TB Guidelines [28]. In all public health facilities, TB diagnostic services and treatment are provided free of charge. Patients initiated on treatment typically receive a one-month supply of medication and are expected to attend monthly clinic visits for drug refills and clinical review.

Community Health Promoters (CHPs), formerly known as Community Health Workers, provide frontline community-based health services. They conduct TB symptom screening, refer suspected cases to health facilities for diagnosis and treatment, and support tracing of patients who miss clinic appointments. CHPs are generally lay community members with basic training who have historically served in voluntary roles, although their roles have increasingly become formalized within the health system. Beyond TB activities, they undertake multiple responsibilities, including health education, maternal and child health support, HIV services, substance abuse awareness, family planning, and nutrition counselling [17]. CHPs operate within Community Health Units (CHUs), each covering approximately 50 households. Kilifi County has an estimated 3,650 CHPs serving an estimated 294,000 households.

### Study Population

For the quantitative component, the study population comprised adult TB patients (aged ≥18 years) who were recorded as lost to follow-up (LTFU) between July 2022 and December 2023, representing the 18 months preceding the study. All TB patients receiving treatment in Kenya are registered and monitored through the national electronic surveillance system, Treatment Information from Basic Unit (TIBU). Patients classified as LTFU were identified through the TIBU system and subsequently linked to their respective Community Health Units (CHUs) and assigned CHPs for tracing. In line with WHO and Kenya National TB Programme guidelines, we adapted the definition of LTFU to include TB patients whose treatment was interrupted for two consecutive months or more and who did not subsequently resume or complete their prescribed treatment course[28].

For the qualitative component, traced TB patients participated in focus group discussions (FGDs). Key informant interviews (KIIs) were conducted with TB clinic in-charges, health service providers, programme managers, CHPs, and members of the County Health Management Teams.

### Data source

TB patients classified as LTFU were identified through the TIBU system and linked to CHPs residing closest to their households. CHPs attempted to establish contact by telephone for patients with recorded mobile numbers and conducted home visits when phone contact was unsuccessful or unavailable. Multiple tracing attempts were made, including repeated household visits and engagement of local administrative leaders where necessary. For patients who could not be traced, information on vital status and whereabouts was obtained from proxy informants, including local administrators, relatives, or neighbours.

For patients who were successfully traced, quantitative data were collected using a structured questionnaire. For the qualitative component, a random sample of traced patients, CHPs, and members of the County Health Management Team participated in in-depth interviews and focus group discussions. Traced patients were referred back to TB clinics, where they received counselling and were restarted on treatment in accordance with Kenya National TB Programme guidelines [29].

### Study size

The quantitative component was descriptive in nature and aimed to characterize TB patients classified as lost to follow-up (LTFU) and subsequently traced. As no formal hypothesis testing was planned, a power calculation was not performed. Instead, the sample size was determined by the number of eligible LTFU patients identified during the study period. Approximately 1,800 TB patients initiate treatment annually in Kilifi County, with an estimated 5% classified as LTFU [30]. Over the 18-month study period, this translated to an expected minimum of approximately 135 LTFU patients eligible for tracing.

For the qualitative component, participant recruitment continued until thematic saturation was achieved. In total, 60 participants were included: 30 successfully traced TB patients participated in five focus group discussions (6 participants per group), and 30 key informants (including CHPs and members of the County Health Management Team) took part in in-depth interviews.

### Data analysis

For the quantitative component, the primary outcome, the proportion of LTFU TB patients successfully traced, was estimated with corresponding exact binomial 95% confidence intervals. As the quantitative variables were categorical, results are presented as frequencies and proportions. Statistical analyses were performed using Stata version 17.0 (StataCorp, College Station, TX, USA).

For the qualitative component, data were analyzed using a thematic analysis approach. Transcripts were reviewed and coded to identify recurrent themes and patterns. The analysis was primarily deductive, guided by the study objectives and the focus on patient and CHP experiences, while allowing for the emergence of additional relevant themes. Data management and coding were conducted using QDA miner lite version 2.0.9 (Provalis research, 2016, https://provalisresearch.com).

### Ethical considerations

All study participants provided written informed consent, and ethical approval was obtained from the AMREF Health Africa Ethics and Scientific Review Committee (ESRC P1523/2023).

## Results

### Quantitative results

Of the 140 TB patients who were LTFU and targeted for tracing, 36/140 (25.7%, 95%CI 18.7‒33.8) were traced. A total of 104/140 (74.3%, 95%CI 66.2‒81.3) patients were not traced, of which 90/140 (64.3%, 95%CI 55.8‒72.2) had moved out of Kilifi County while 14/140 (10%, 95%CI 5.6‒16.2) had died (**Fig 1**).

**Fig 1.**
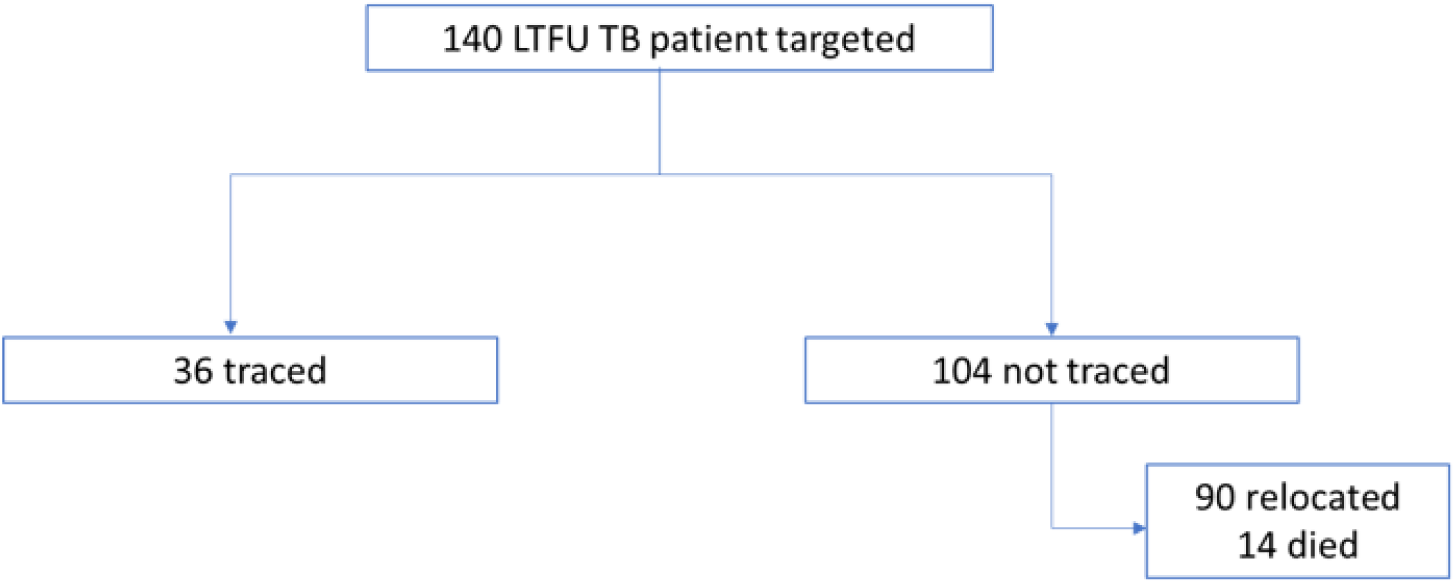
Study participants flow chart.

Of the 36 LTFU patients traced, one-quarter were less than 25 years old while 14 (38.9%) were >35 years old. The ratio of male: female was 1:1. More than three-quarters were from Kilifi North and south sub-counties. Only 2 (5.6%) had never received any formal education while 11 (30.6%) were unemployed. Three (8.3%) were HIV infected while 1 (2.8%) had cancer, the rest (n=32, 88.9%) had no underlying medical conditions. A total of 25 (69.4%) were currently not using any recreation drugs as shown in **Table 1** below.

**Table 1.**
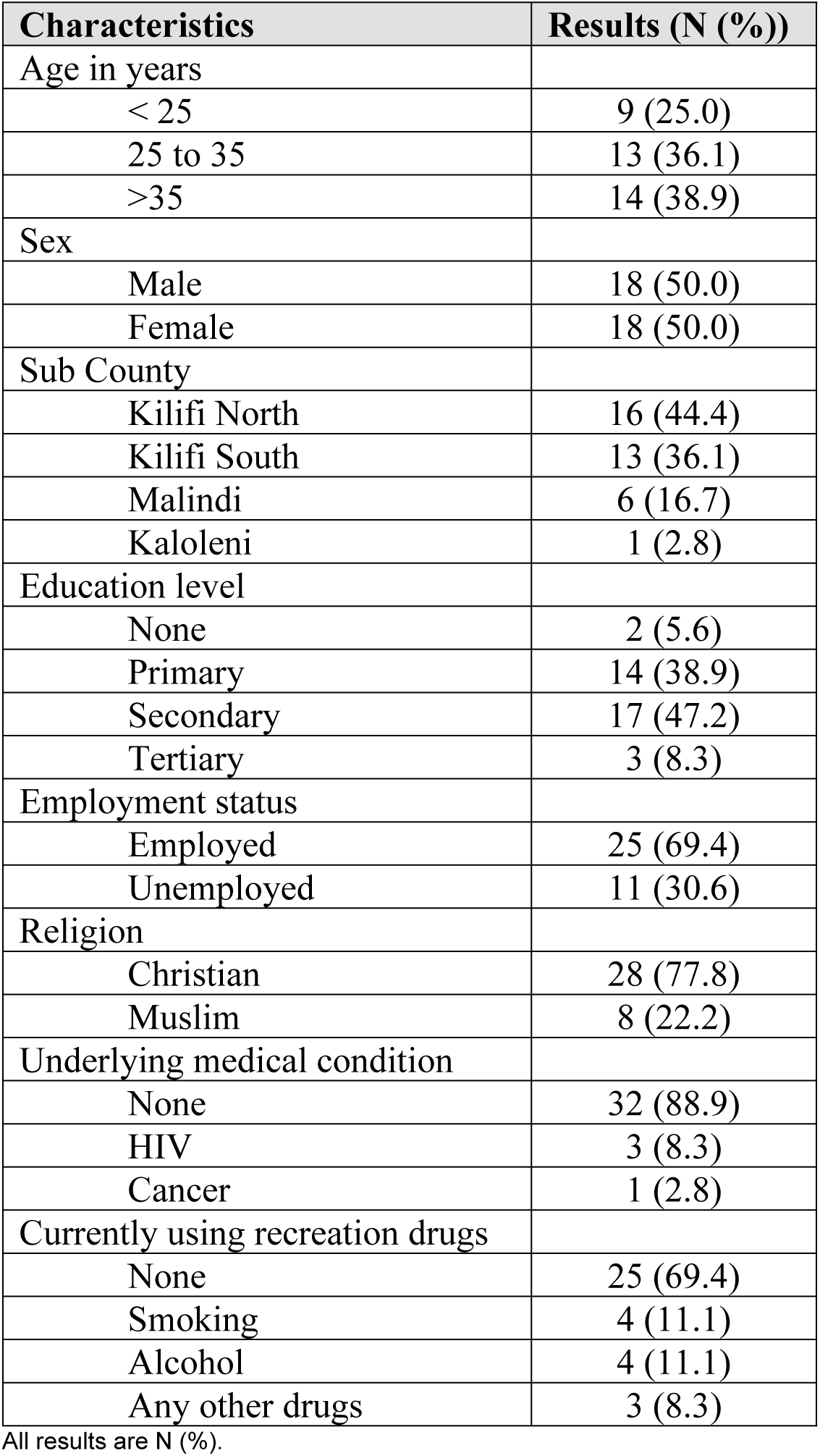
Characteristics of LTFU patients traced.

| Characteristics | Results (N (%)) |
| --- | --- |
| Age in years |  |
| < 25 | 9 (25.0) |
| 25 to 35 | 13 (36.1) |
| >35 | 14 (38.9) |
| Sex |  |
| Male | 18 (50.0) |
| Female | 18 (50.0) |
| Sub County |  |
| Kilifi North | 16 (44.4) |
| Kilifi South | 13 (36.1) |
| Malindi | 6 (16.7) |
| Kaloleni | 1 (2.8) |
| Education level |  |
| None | 2 (5.6) |
| Primary | 14 (38.9) |
| Secondary | 17 (47.2) |
| Tertiary | 3 (8.3) |
| Employment status |  |
| Employed | 25 (69.4) |
| Unemployed | 11 (30.6) |
| Religion |  |
| Christian | 28 (77.8) |
| Muslim | 8 (22.2) |
| Underlying medical condition |  |
| None | 32 (88.9) |
| HIV | 3 (8.3) |
| Cancer | 1 (2.8) |
| Currently using recreation drugs |  |
| None | 25 (69.4) |
| Smoking | 4 (11.1) |
| Alcohol | 4 (11.1) |
| Any other drugs | 3 (8.3) |

All results are N (%).

More than one-half of the traced patients were the household heads (n=20, 55.6%). Among the 16 patients who were not household head, 14/16 (87.5%) reported the household head was away when they were diagnosed with TB while 13/16 (81.3%) required permission from the household head to attend TB clinic. There were 7 (19.4%) patients who lived in households with more than five members. All the 36 patients either had access (n=12, 33.3%) or owned (n=24, 66.7%) a mobile phone. A total of 32 (88.9%) were using solid fuels (wood/charcoal) as source of energy for cooking. Eight (22.2%) patients walked to hospital, of which 6/8 (75%) took 30mins to one hour walking to hospital and 2/8 (25%) took two to four hours walking to hospital (**Table 2**).

**Table 2.** Household characteristics of traced patients.

| Characteristics | Results (N (%)) |
| --- | --- |
| Who is the head of the household? |  |
| Self | 20 (55.6) |
| Spouse | 8 (22.2) |
| Parent | 8 (22.2) |
| Was the head of family aware you were diagnosed with TB? (N=16) |  |
| No | 2 (12.5) |
| Yes | 14 (87.5) |
| Do you require permission from household head to attend TB clinic? |  |
| No | 3 (18.8) |
| Yes | 13 (81.3) |
| Type of house |  |
| Mud wall | 14 (38.9) |
| Stone wall | 22 (61.1) |
| How many people live in the house? |  |
| < 5 members | 29 (80.6) |
| ≥5 members | 7 (19.4) |
| Water source |  |
| Tap | 31 (86.1) |
| Borehole/well in the community | 5 (13.9) |
| Toilet type |  |
| Shared toilet in compound | 26 (72.2) |
| Toilet in house | 9 (25.0) |
| None/Flying | 1 (2.8) |
| Do you own a mobile phone? |  |
| Own one | 24 (66.7) |
| Access to one | 12 (33.3) |
| Source of fuel for household cooking |  |
| Wood or charcoal | 32 (88.9) |
| LPG gas | 4 (11.1) |
| Means of transport to hospital |  |
| Walking | 8 (22.2) |
| Motorcycle | 27 (75.0) |
| Public vehicle | 1 (2.8) |
| Walking time to hospital |  |
| 30 mins to one hour | 6 (75.0) |
| Two to four hours | 2 (25.0) |

All results are N (%).

Four (11.1%) of the traced patient reported someone within the family had been diagnosed with TB before them, of which 1/4 (25%) did not complete TB treatment as well. Less than one-half of the traced patients had been linked with a DOT provider (n=16, 44.4%). Among the 16 patients linked to a DOT provider, 15/16 (93.75) reported that the DOT provider was supportive in TB treatment. Three quarters reported their spouse/family members were supportive in TB treatment. A total of 27/36 (75%) of the traced patients were willing to restart TB treatment. One traced patient recommended DOT providers to bring drugs to community because of distance (**Table 3**)

**Table 3.** History of TB treatment of TB traced patients.

| Characteristics | Results (N %) |
| --- | --- |
| Has anyone in the family been diagnosed with TB before you? |  |
| No | 32 (88.9) |
| Yes | 4 (11.1) |
| If family history of TB, did they complete TB treatment (N=4) |  |
| No | 1 (25.0) |
| Yes | 3 (75.0) |
| Were you linked with a DOT provider? |  |
| No | 20 (55.6) |
| Yes | 16 (44.4) |
| Was the DOT provider supportive in TB treatment? (N=16) |  |
| No | 1 (6.3) |
| Yes | 15 (93.7) |
| Was your spouse and family members supportive in TB treatment? |  |
| No | 9 (25.0) |
| Yes | 27 (75.0) |
| Are you willing to restart TB treatment |  |
| No | 9 (25.0) |
| Yes | 27 (75.0) |
| Recommended changes in TB management to improve retention in care |  |
| None | 32 (88.9) |
| DOTs providers to bring drugs to community because of distance | 1 (2.8) |
| Prefer injections to reduce drug burden | 1 (2.8) |
| More follow-ups for TB patients | 1 (2.8) |
| Quality service provision | 1 (2.8) |

All results are N (%).

The leading reported reason for stopping taking TB drugs was that the patients felt disease symptoms had resolved (n=16, 44.4% (27.9‒61.9). A total of 10 patients (27.8%, 14.2‒45.2) stopped taking TB drugs because of cost of travelling to pick drugs. A similar proportion (27.8%, 14.2‒45.2) stopped taking TB drugs because of drugs side effects.

### Qualitative results

**Table 4** summarizes the qualitative findings highlighting the major themes and illustrative participant perspectives.

**Table 4:**
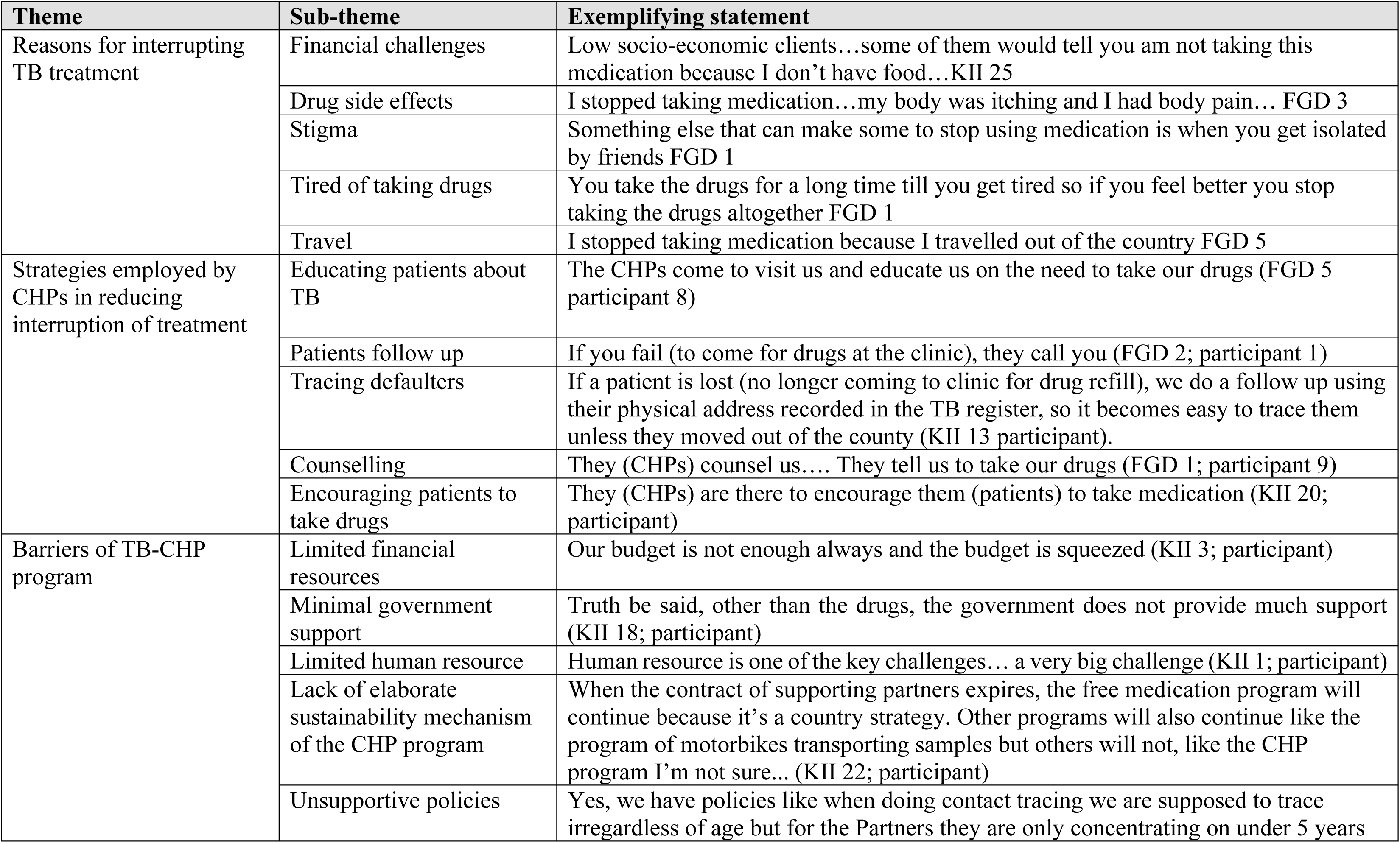
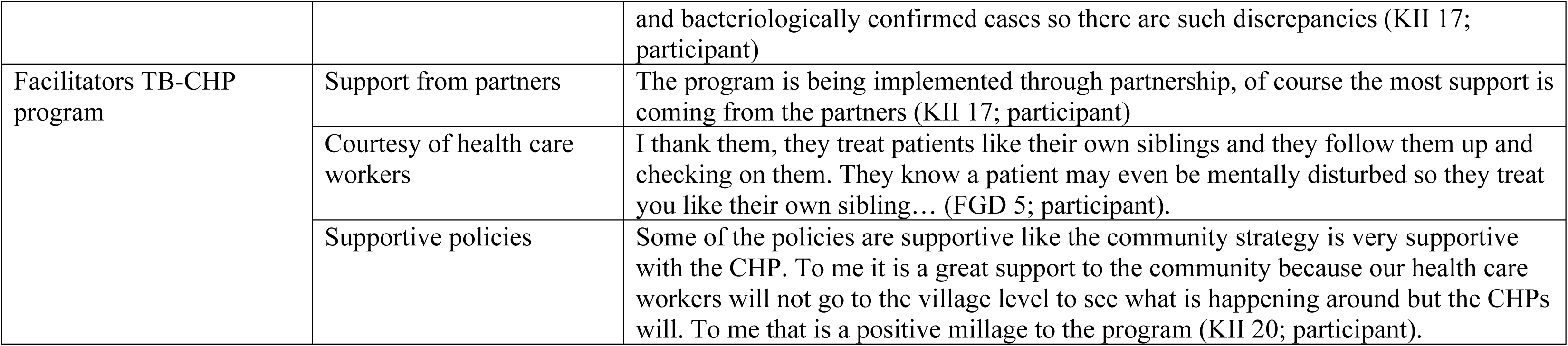
Summary of qualitative findings illustrating themes, sub themes and participants exemplifying statements.

### Reasons for interrupting TB treatment

Traced TB patients discussed several reasons for interrupting TB treatment, including financial hardship, medication side effects, stigma, treatment fatigue, and travel. Discussions highlighted how lack of food and financial constraints could make it difficult to continue treatment, while adverse effects and feeling better after prolonged treatment could lead some patients to stop taking their medication. Stigma and social isolation were also identified as barriers to treatment continuity, while travel disrupted treatment for some patients.

### Strategies used by Community Health Promoters

Discussions highlighted several strategies used by CHPs to support treatment continuity, including patient education, counselling, encouragement to adhere to treatment, follow-up of missed clinic appointments, and tracing patients who interrupt treatment. CHPs used contact information and physical addresses recorded in TB registers to locate patients who missed appointments, although tracing was more difficult when patients had moved outside the county.

### Barriers to implementation of the TB–CHP program

Despite positive perceptions of the program, interviews with KIIs identified several barriers to implementing the CHP programme, including inadequate financial resources, limited government support, shortages of human resources, and the absence of clear mechanisms for sustaining the programme beyond partner funding. Participants also highlighted policy inconsistencies, particularly differences between government and partner priorities for contact tracing, which limited the scope and continuity of CHP-led activities.

### Facilitators of the TB–CHP program

Key facilitators of the TB–CHP program included strong support from implementing partners, compassionate and supportive interactions between healthcare workers and patients, and policies that promote community-based health services. The community health strategy was viewed as particularly valuable in extending healthcare services to the village level, enabling CHPs to reach patients in their communities and provide follow-up and support that health facility staff may not be able to provide (**Table 4**).

## Discussion

In this setting, we were only able to trace one-quarter of TB patients who had discontinued treatment and were classified as LTFU. Although guidelines recommend systematic monitoring of TB treatment outcomes, active tracing of patients who interrupt treatment is not routinely or effectively implemented in our context. Engaging CHPs, who reside within the same communities as patients, represents a pragmatic and potentially low-cost strategy to re-engage individuals in care. However, our findings highlight an important limitation of community-based tracing strategies, the ability of CHPs to re-engage patients depends not only on the availability and commitment of community health workers, but also on the accuracy of patient contact information and the mobility and social circumstances of patients. This also underscores the importance of strengthening inter-county coordination and formal patient transfer systems to ensure continuity of care, rather than relying on informal self-referral. We were unable to ascertain whether patients who relocated continued TB treatment at their new locations, highlighting gaps in patient tracking and national surveillance systems. This is particularly important for TB programmes, where treatment interruption can increase the risk of poor treatment outcomes and ongoing transmission. Previous studies have shown that when tracing is initiated promptly after a missed visit or LTFU event, up to 55% of patients can be successfully traced and restarted on treatment [31]. Our lower tracing yield may therefore reflect delays in tracing or population mobility in this setting. Additionally, the identification of deaths among individuals initially recorded as LTFU suggests that TB programs may underestimate mortality when outcomes among LTFU patients are not actively verified.

In Kenya, TB diagnosis and treatment in public health facilities are provided free of charge. However, government support primarily covers the direct costs of diagnostic tests and anti-TB medications, leaving substantial indirect costs to the patient. Both patients and healthcare providers perceive this support as insufficient to sustain successful treatment completion. Most patients in our setting are from low socioeconomic backgrounds and struggle to afford transportation to health facilities for drug refills, as well as adequate daily nutrition, factors that contributed to treatment interruption. Economic hardship is widely recognized as a major barrier to ending TB, and the WHO End TB Strategy calls for eliminating catastrophic costs for TB-affected households [32–34]. Beyond medical expenses, patients often face indirect costs such as transport, food, and lost income due to time away from work, all of which can deplete household resources and negatively affect treatment adherence and outcomes [35, 36].

The quantitative findings reinforced by the qualitative discussions suggest that treatment interruption was driven by a combination of individual, social, and health-system factors. The most common reason for stopping treatment was feeling better or perceiving that TB symptoms had resolved, followed by the cost of travelling to collect medication and medication side effects. Stigma, a well-established social determinant of health also emerged as an important contributor to LTFU[37]. Strengthening psychosocial support from family members, communities, and healthcare providers may therefore improve treatment retention [38]. Although drug side effects were reported as a reason for treatment discontinuation, standard six-month first-line TB regimens are generally well tolerated [39]. In our setting, routine laboratory monitoring for drug toxicity is not systematically performed unless clinically indicated, which may contribute to patient concerns about adverse effects and influence adherence. Together, these findings suggest that LTFU should not be viewed solely as a failure of individual adherence. Rather, treatment interruption occurs within a broader context in which patients may face substantial economic, social, and practical barriers to completing a prolonged course of treatment. Interventions to reduce LTFU therefore need to address both patient-level barriers and the structural conditions that make sustained treatment adherence difficult.

CHPs serve as the first point of contact for many community members seeking health services, providing health education, basic care, and linkage to formal health facilities. In Kenya, CHPs have historically worked on a voluntary basis, with limited and often inconsistent financial support from implementing partners such as USAID and other non-governmental organizations. In the context of TB care, CHPs are expected to support patients throughout treatment by providing education, counselling, adherence support, and tracing individuals who miss scheduled clinic visits, in addition to fulfilling their broader community health responsibilities [17, 22]. However, our qualitative findings suggest that CHPs were few in number and inadequately supported financially to conduct active and sustained TB patient tracing. Their workload is substantial, with competing priorities that often emphasize services for children under five years, pregnant women, and bacteriologically confirmed TB cases [22]. Despite these challenges, CHPs are generally well respected and positively perceived within communities, and their guidance and referrals to the formal health system are highly valued by patients with TB, HIV, and other chronic conditions [21].

While CHPs have considerable potential to strengthen TB treatment continuity through patient tracing, their effectiveness is constrained by limited staffing and chronic underfunding. Reliance on short-term implementing partner support is neither sustainable nor equitable, as it depends on partners’ geographic focus, programmatic priorities, and funding cycles, and may create uncertainty and low morale among CHPs. Strengthening government investment and formal integration of CHPs into TB programs is therefore critical to optimize their contribution to TB control efforts. CHPs balance professional responsibilities with family and community commitments, underscoring the need for sustainable remuneration [40, 41]. In 2024, the Government of Kenya recruited approximately 105,000 CHPs and introduced a monthly stipend of KES 5,000 (approximately USD 40), jointly funded by the national and county governments. This represents an important step toward formal recognition and support of the community health workforce. However, financial stipends alone are insufficient. CHPs also require consistent provision of supplies, structured training, supportive supervision, and ongoing mentorship to effectively fulfil their roles and contribute meaningfully to TB control and other community health priorities.

A key strength of this study is the use of both quantitative and qualitative data, which enabled us to capture experiences and perspectives directly from TB patients and CHPs. However, the study has several limitations. First, the study was conducted in Kilifi County, and the findings may not be generalisable to other counties or settings with different TB epidemiology, population mobility, or community health structures. We included only TB patients who were lost to follow-up (LTFU) in 2023 and the CHPs responsible for their community health units, excluding CHPs from other CHUs and patients who completed treatment. The experiences of these excluded groups may differ from those included in the study. In addition, the study was not powered to detect statistically significant differences, as the sample size was determined by the number of LTFU patients eligible for tracing rather than formal sample size calculations. Despite these limitations, our findings are consistent with previous research, reinforcing the relevance of our observations [21, 40, 41].

## Conclusion

CHP-led tracing offers an important opportunity to strengthen continuity of TB care in Kenya, but its impact is constrained by patient mobility and limited programme resources. Strengthening and sustainably financing CHPs, improving cross-county patient tracking, and addressing the financial, social, and treatment-related barriers to adherence are critical to preventing treatment interruption and improving TB treatment outcomes.

## Data Availability

The study anonymized data underlying the results presented are available in the supporting materials (S1 Table).

## Acknowledgement

We sincerely thank all the study participants, CHPs involved and the support from the Kilifi County TB program staff. We also acknowledge the contributions of the study team members and data management personnel whose dedication made this work possible.

This work was supported by Global Fund, Amref Health Africa in Kenya (Grant Number: Ken-T-Amref). The views expressed in this publication are those of the authors and do not necessarily reflect the views of the funding agency.

## Author contributions

**Conceptualization:** Moses M. Ngari, Stevenson K. Chea, Geoffrey Katana, Osman A. Abdullahi

**Data curation:** Deche Sanga, Fredrick M. Thuva, George U. Onyango, Stevenson K. Chea, Geoffrey Katana

**Formal analysis:** Moses M. Ngari, Stevenson K. Chea

**Investigation:** Geoffrey Katana, Deche Sanga, Stevenson K. Chea

**Methodology:** Moses M. Ngari, Deche Sanga, Stevenson K. Chea, Geoffrey Katana, Osman A. Abdullahi.

**Project administration:** Christine M. Mwamsidu, Catherine Kamau, Fredrick M. Thuva

**Supervision:** Osman A. Abdullahi

**Validation:** Osman A. Abdullahi

**Visualization:** Moses M. Ngari, Stevenson K. Chea

**Writing-original draft:** Moses M. Ngari, Stevenson K. Chea

**Writing-review & editing:** Moses M. Ngari, Stevenson K. Chea, Deche Sanga, Geoffrey Katana, Christine M. Mwamsidu, Catherine Kamau, Fredrick M. Thuva, Osman A. Abdullahi.

